# Estimation of disease burden and clinical severity of COVID-19 caused by Omicron BA.2 in Shanghai, February-June 2022

**DOI:** 10.1101/2022.07.11.22277504

**Authors:** Xinhua Chen, Xuemei Yan, Kaiyuan Sun, Nan Zheng, Ruijia Sun, Jiaxin Zhou, Xiaowei Deng, Tingyu Zhuang, Jun Cai, Juanjuan Zhang, Marco Ajelli, Hongjie Yu

## Abstract

**Background:** An outbreak of COVID-19 caused by the SARS-CoV-2 Omicron BA.2 sublineage occurred in Shanghai, China from February to June 2022. The government organized multiple rounds of molecular test screenings for the entire population, providing a unique opportunity to capture the majority of subclinical infections and better characterize disease burden and the full spectrum of Omicron BA.2 clinical severity.

**Methods:** Using daily reports from the websites of the Shanghai Municipal Health Commission, we estimated the incidence of infections, severe/critical infections, and deaths to assess the disease burden. By adjusting for right censoring and Reverse Transcription-Polymerase Chain Reaction (RT□PCR) sensitivity, we provide estimates of clinical severity, including the infection fatality risk, symptomatic case fatality risk, and risk of developing severe/critical disease upon infection.

**Findings:** From February 26 to June 30, 2022, the overall infection rate, severe/critical infection rate, and mortality rate were 2.74 (95% CI: 2.73-2.74) per 100 individuals, 6.34 (95% CI: 6.02-6.66) per 100,000 individuals and 2.42 (95% CI: 2.23-2.62) per 100,000 individuals, respectively. The severe/critical infection rate and mortality rate increased with age with the highest rates of 125.29 (95% CI: 117.05-133.44) per 100,000 and 57.17 (95% CI: 51.63-62.71) per 100,000 individuals, respectively, noted in individuals aged 80 years or older. The overall fatality risk and risk of developing severe/critical disease upon infection were 0.09% (95% CI: 0.08-0.10%) and 0.23% (95% CI: 0.20-0.25%), respectively. Having received at least one vaccine dose led to a 10-fold reduction in the risk of death for infected individuals aged 80 years or older.

**Interpretation:** Under the repeated population-based screenings and strict intervention policies implemented in Shanghai, our results found a lower disease burden and mortality of the outbreak compared to other settings and countries, showing the impact of the successful outbreak containment in Shanghai. The estimated low clinical severity of this Omicron BA.2 epidemic in Shanghai highlight the key contribution of vaccination and availability of hospital beds to reduce the risk of death.

**Funding:** Key Program of the National Natural Science Foundation of China (82130093).

**Research in context:** *Evidence before this study:* We searched PubMed and Europe PMC for manuscripts published or posted on preprint servers after January 1, 2022 using the following query: (“SARS-CoV-2 Omicron”) AND (“burden” OR “severity”). No studies that characterized the whole profile of disease burden and clinical severity during the Shanghai Omicron outbreak were found. One study estimated confirmed case fatality risk between different COVID-19 waves in Hong Kong; other outcomes, such as fatality risk and risk of developing severe/critical illness upon infection, were not estimated. One study based on 21 hospitals across the United States focused on Omicron-specific in-hospital mortality based on a limited sample of inpatients (565). In southern California, United States, a study recruited more than 200 thousand Omicron-infected individuals and estimated the 30-day risk of hospital admission, intensive care unit admission, mechanical ventilation, and death. None of these studies estimated infection and mortality rates or other indictors associated with disease burden. Overall, the disease burden and clinical severity of the Omicron BA.2 variant have not been fully characterized, especially in populations predominantly immunized with inactivated vaccines.

*Added value of this study:* The large-scale and multiround molecular test screenings conducted on the entire population during the Omicron BA.2 outbreak in Shanghai, leading to a high infection ascertainment ratio, provide a unique opportunity to capture the majority of subclinical infections. As such, our study provides a comprehensive assessment of both the disease burden and clinical severity of the SARS-CoV-2 Omicron BA.2 sublineage, which are especially lacking for populations predominantly immunized with inactivated vaccines.

*Implications of all the available evidence:* We estimated the disease burden and clinical severity of the Omicron BA.2 outbreak in Shanghai in February-June 2022. These estimates are key to properly interpreting field evidence and assessing the actual spread of Omicron in other settings. Our results also provide support for the importance of strategies to prevent overwhelming the health care system and increasing vaccine coverage to reduce mortality.

## Background

An outbreak of coronavirus disease 2019 (COVID-19) caused by the severe acute respiratory syndrome coronavirus 2 (SARS-CoV-2) Omicron BA.2 sublineage has occurred in Shanghai, China from February to June 2022. To contain the outbreak in the shortest period of time, the Shanghai government has implemented a set of strict nonpharmacological interventions (NPIs). Between March 16 and March 27, Shanghai adopted grid management at the subdistrict level, fractioning subdistricts into high-risk areas and nonhigh-risk areas according to the number of cases and infections in the area. In addition, several rounds of population-wide nucleic acid test screenings were performed within high-risk areas and nonhigh-risk areas. However, such interventions were not sufficient to control community spread. The entire city was forced to enter a phased stage of lockdowns starting on March 28 when Shanghai’s Pudong district entered a population-wide lockdown, and the entire city entered a lockdown phase on April 1, 2022. The epidemic reached its inflection point on April 13, 2022, and the outbreak was eventually brought under control. On June 1, 2022, the Shanghai government declared the end of the city-wide lockdown. At that time, the total number of reported infections was 626,811, including 568,811 asymptomatic infections, 58,000 symptomatic cases, and 588 deaths.^1^

One of the reasons that prompted Shanghai to adopt stringent NPIs was the vaccination gap in Shanghai’s elderly population. In fact, as of April 15, 2022, only 62% and 38% of people older than 60 years old completed primary and booster vaccinations, respectively.^2^ The generally low coverage of primary and booster vaccination among the elderly – the population at highest risk of severe and critical COVID-19 disease – could have led to a large number of deaths if the outbreak were left to spread uncontrolled. In comparison, an uncontained Omicron outbreak in Hong Kong, China resulted in 9,188 deaths between Dec 31, 2021 and June 30, 2022.^3^

Estimation of disease burden and clinical severity is critical to identify appropriate intervention strategies, plan for health care needs, and ensure that the health system operates properly. In particular, the disease burden and clinical severity of the Omicron BA.2 variant have not been fully characterized, especially in populations predominantly immunized with inactivated vaccines. Furthermore, during this outbreak in Shanghai, the government organized multiple rounds of molecular test screenings for the entire population, which led to a high infection ascertainment ratio, thus providing a unique opportunity to capture the majority of subclinical infections and better characterize the full spectrum of Omicron BA.2 clinical severity.

In this study, we used publicly reported data from the Shanghai Municipal Health Commission to estimate the incidence of infections, severe/critical infections, and deaths to assess the disease burden of the Omicron BA.2 outbreak in Shanghai in early 2022. In addition, we provide estimates of the infection fatality risk and symptomatic case fatality risk as well as the risk of developing severe/critical disease upon infection to assess the clinical severity of Omicron BA.2 during the outbreak.

## Methods

### Case definition

The definition of confirmed COVID-19 cases and SARS-CoV-2 infection was based on the Clinical Guidance for COVID-19 Pneumonia Diagnosis and Treatment (trial ninth edition) and the COVID-19 Prevention and Control Protocol (eighth edition) published by the National Health Commission (NHC) of China.^4,5^ SARS-CoV-2 infections, including asymptomatic infections and symptomatic cases, were ascertained by Reverse Transcription-Polymerase Chain Reaction (RT□PCR). Symptomatic cases were further categorized by clinical severity into mild, moderate, severe, and critical cases. Mild cases were defined as having mild symptoms, such as fever, fatigue, loss of taste/smell but without radiographic evidence of pneumonia. Moderate cases were those with typical symptoms of a respiratory infection (e.g., fever, dry cough, fatigue, etc.) and radiographic evidence of pneumonia. Severe cases referred to those patients with at least one of the following conditions: breathing problems, low oxygen saturation, low PaO_2_/FiO_2_ (PaO_2_ denotes partial pressure of oxygen in arterial blood; FiO_2_ denotes fraction of inspired oxygen), or progressive symptoms combined with pulmonary imaging showing obvious progress of lesions (>50%) within 24-48 hours. Critical cases referred to patients who met any one of the following three criteria: respiratory failure, shock, or organ failure that required intensive care unit admission. Asymptomatic infections were defined as RT□PCR-positive individuals who did not meet any of the following clinical criteria: 1) fever, cough, sore throat, and other self-perceived and clinically identifiable symptoms or signs; 2) no radiographic evidence of pneumonia.

### Case identification and surveillance

During the Omicron BA.2 outbreak in Shanghai, multiple mass population screenings using RT□PCR tests were conducted for the entire city population to identify infected individuals, including asymptomatic and presymptomatic individuals **(Figure S1)**.^6,7^ Moreover, self-performed rapid antigen test screenings of the population were performed as a supplement of nucleic acid tests; any positive result of an antigen test required confirmation by a nucleic acid test. Routine surveillance was mainly based on symptom-based surveillance from medical institutions. Contact tracing, epidemiological investigation, and screening of high-risk populations were also conducted.

### Data sources and parameters

Daily reports on the cumulative number of infections, symptomatic cases, and deaths were extracted from the websites of the Shanghai Municipal Health Commission since February 26, 2022, which represents the beginning of the Omicron BA.2 outbreak in Shanghai.^1^ The distribution of age and vaccination status of infections, severe/critical patients, and deaths before May 13, 2022 were extracted from the literature (*Wang et al*., *personal communication*). The data of deceased cases after May 13, 2022 were extracted from the report of Shanghai Municipal Health Commissions.^1^ All data sources and parameters used are listed in **Table S1**.

### Primary outcomes

To assess the burden of the investigated outbreak, we estimated the prevalence of infections, cases of severe/critical disease, and deaths. To assess the clinical severity of the Omicron BA.2 variant during the investigated outbreak, we estimated the proportion of asymptomatic infections (P_asym_), infection fatality risk (IFR), risk of severe/critical disease given the infection (ISR), symptomatic case-fatality risk (sCFR), and risk of severe/critical disease given a symptomatic infection (sCSR). Specifically, the P_asym_ was defined as the ratio between the number of asymptomatic infections and the total number of infections. The IFR was defined as the ratio between the number of deaths and the total number of SARS-CoV-2 infections, which includes both asymptomatic infections and symptomatic cases confirmed by RT□PCR. ISR was defined as the ratio between the number of severe/critical cases and the total number of SARS-CoV-2 infections confirmed by RT□PCR. The sCFR was defined as the ratio between the number of deaths and the number of symptomatic COVID-19 cases confirmed by RT□PCR. The sCSR was defined as the ratio between the number of severe/critical cases and the number of symptomatic COVID-19 cases confirmed by RT□PCR **(Figure 1)**.

**Figure 1.**
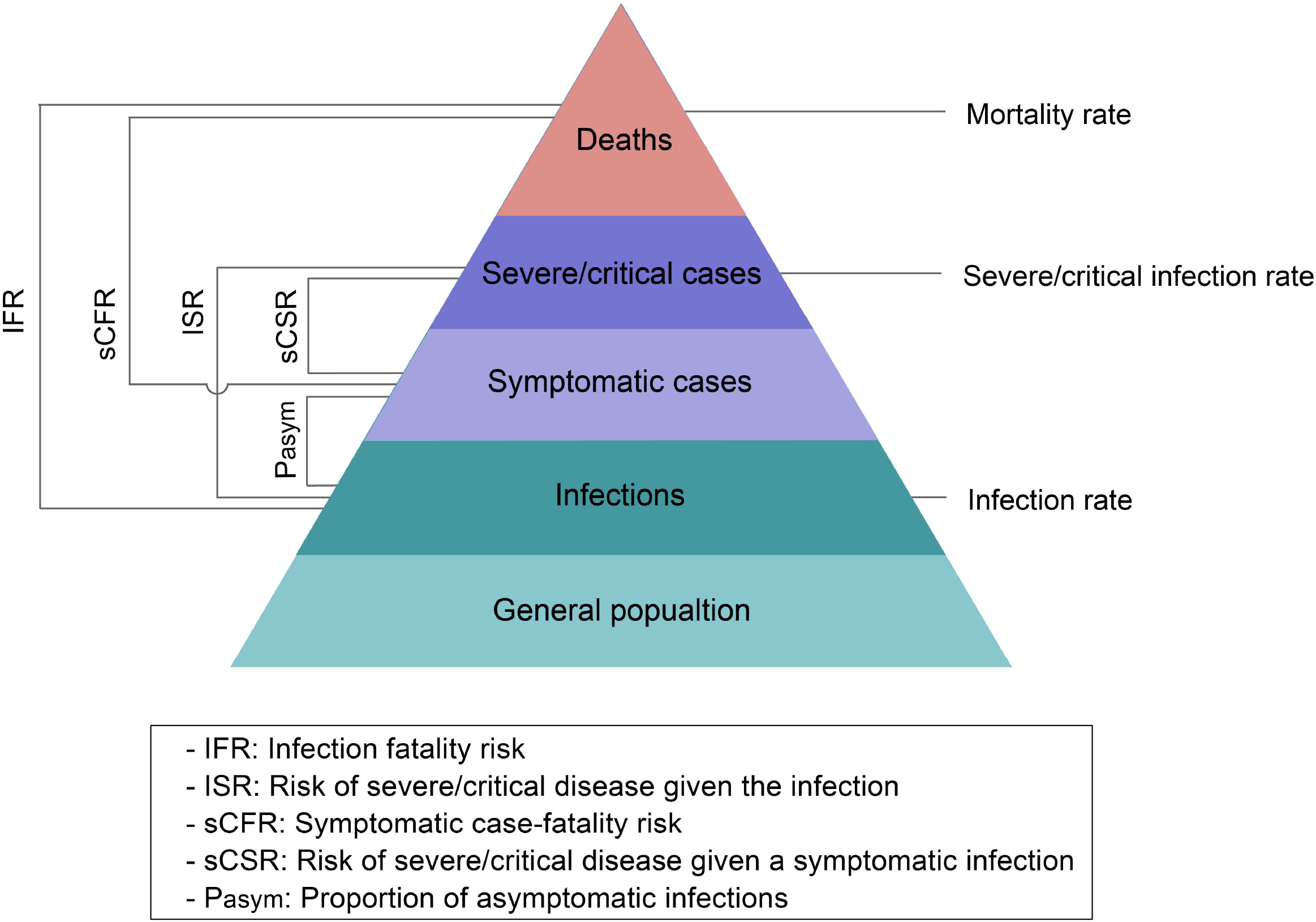
Spectrum of COVID-19 and primary outcomes.

### Statistical analysis

#### Estimation of the disease burden

Given that the cumulative number of severe/critical cases at the end of the outbreak was not officially reported, we extracted the number of severe/critical cases before May 13 from the literature (*Wang et al*., *personal communication*). Based on the assumption that the risk of death among severe/critical patients remained constant from May 13 until the end of the outbreak, we used the age-specific risk to estimate the cumulative number of severe/critical infections given the cumulative number of deaths by age category. We estimated a set of age-specific attack rates between February 26 and June 30, 2022 by dividing the total number of reported infections, severe/critical patients, and deaths by the population in each age group.

#### Estimation of clinical severity

We estimated both crude and adjusted P_asym_, IFR, ISR, sCFR, and sCSR to assess the clinical severity of Omicron BA.2 during the Shanghai outbreak. Using Garkse’s method,^8^ we adjusted for right censoring by weighting the denominator with the distribution of the time interval from infection to symptom onset, infection to severe/critical illness, infection to death, symptom onset to severe/critical illness, and symptom onset to death (**Equations 1-5**).

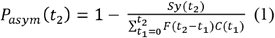

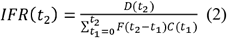

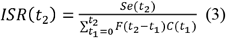

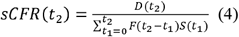

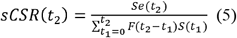

where the numerator refers to the cumulative number of cases with predefined endpoints on Day *t*_2_ (*Sy*: symptomatic infection; *Se*: severe and critical infection; *D*: death), and the denominator refers to a weighted sum of daily (*t*_1_) reported total number of confirmed infections **(Equations 1-3)** or symptomatic cases **(Equations 4-5)**. The weights are based on the density distribution of the time interval from infection to symptom onset, from infection to severe/critical illness, from infection to death, from symptom onset to severe/critical illness, and from symptom onset to death (F). Essentially, this design allows the exclusion of a proportion of recent cases for whom the final outcome has yet to be observed at the early stage of outbreak. To obtain the interval parameter from infection to death, we resampled three intervals (interval from infection to symptom onset; onset to admission; admission to death) according to studies that reported estimated distributions of key time to event of Omicron BA.2 (*Yu et al*., *unpublished data*). Then, we add these three intervals for each individual category and fit gamma, lognormal, and Weibull distributions to identify the best estimates according to the Akaike Information Criterion (AIC) and the Bayesian Information Criterion (BIC). We used a similar method to estimate the interval from infection to severe/critical illness. Additionally, we further adjusted the number of total infections by randomly simulating 10,000 draws of RT□PCR sensitivity from a binomial distribution to test the uncertainty of the parameter regulating the RT□PCR sensitivity. We conducted a sensitivity analysis by changing the parameter of another widely used RT□PCR kit in Shanghai and re-estimated the burden and clinical severity **(Figure S4-S5)**. We further estimated the IFR and ISR stratified by age groups (3-17, 18-39, 40-59, 60-79, and ≥ 80 years) and vaccination status (not vaccinated and vaccinated). People who received at least one dose of vaccine were defined as ‘vaccinated’; otherwise, they were defined as ‘unvaccinated’. Binomial distributions were used to estimate 95% CIs.

### Role of the funding source

The funder of the study had no role in the study design, data collection, data analysis, data interpretation, or writing of the report. The corresponding authors had full access to all the data in the study and had final responsibility for the decision to submit for publication.

This study was approved by the institutional review board of the School of Public Health, Fudan University (IRB# 2022-05-0968). All data were collected from publicly available sources. Data were deidentified, and the need for informed consent was waived.

## Results

From February 26 to June 30, 2022, a total of 627,115 confirmed infections were officially reported, including 58,137 symptomatic cases, 568,978 asymptomatic infections, and 588 deceased cases. The overall infection rate was 2.74 (95% CI: 2.73-2.74) per 100 individuals. The lowest infection rate was noted in the population aged 3-17 years (1.67, 95% CI: 1.66-1.69 per 100 individuals), whereas the highest rate in the population aged 60-79 years (3.65, 95% CI: 3.63-3.67 per 100 individuals). **(Figure 2A)**. The rate of severe/critical infection increased with age. The highest rate of 125.29 (95% CI: 117.05-133.44) per 100,000 individuals was noted in individuals aged 80 years or older, and the lowest rate was observed in individuals aged 3-17 years (<0.001 per 100,000 individuals) **(Figure 2B)**. The overall mortality rate was 2.42 (95% CI: 2.23-2.62) per 100,000 individuals. Regarding specific age groups, the mortality rate was 0 in 3- to 17-year-old individuals, 0.02 (95% CI: 0-0.04) per 100,000 individuals in those aged 18-39 years, 0.32 (95% CI: 0.19-0.45) per 100,000 individuals in those aged 40-59 years, 4.60 (95% CI: 3.87-5.32) per 100,000 individuals in those aged 60-79 years, and 57.17 (95% CI: 51.63-62.71) per 100,000 individuals in those aged 80 years or older **(Figure 2C)**.

**Figure 2.**
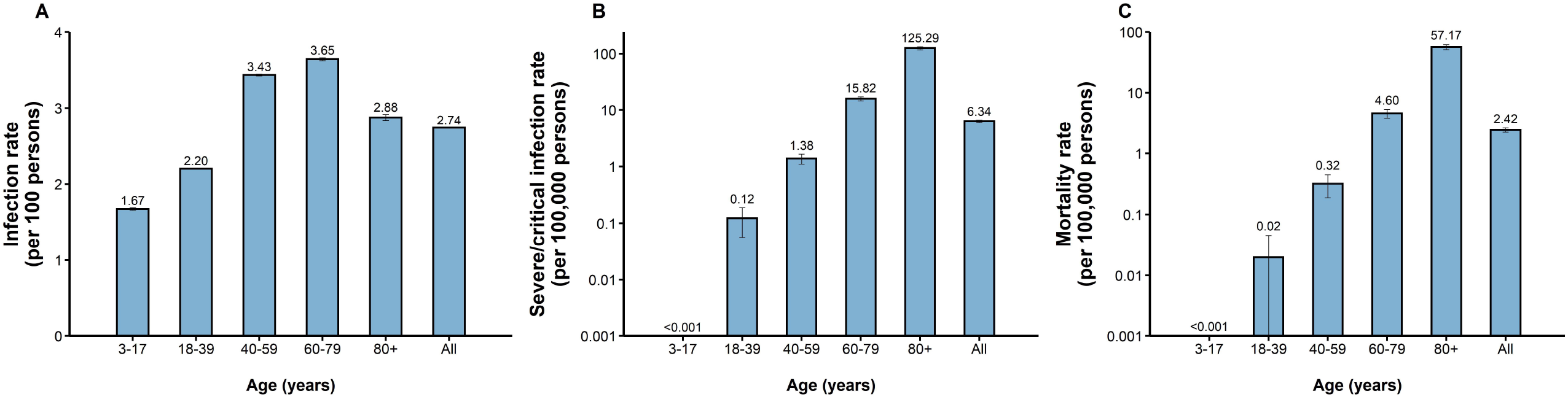
Infection rate, severe/critical infection rate and mortality rate of COVID-19 caused by SARS-CoV-2 Omicron in Shanghai. **A)** Infection rate (per 100 individuals); **B)** Severe/critical infection rate (per 100,000 individuals); **C)** Mortality rate (per 100,000 individuals)

By June 30, 2022, the adjusted P_asym_ was 90.7% (95% CI: 90. 7-90.8%) **(Figure S2)**. The overall adjusted IFR was 0.09% (95% CI: 0.08-0.09%) with an extremely low risk of fatality for infected individuals younger than 60 years. The adjusted IFR was 0.13% (95% CI: 0.11-0.14%) for individuals aged 60-79 and 1.99% (95% CI: 1.76-2.11%) for those aged 80 years or older **(Figure 3A)**. The adjusted IFR for unvaccinated infected individuals aged 80 years or older was 2.22% (95% CI: 1.96-2.35%), which was approximately 10-fold higher than that of those who had received at least one COVID-19 dose (0.25%, 95% CI: 0.21-0.25%). The adjusted IFR for individuals aged 60-79 years was 0.02% (95% CI: 0.02-0.03%) for vaccinated individuals and 0.31% (95% CI: 0.27-0.33%) for unvaccinated individuals **(Figure 3A)**. The overall adjusted ISR was 0.23% (95% CI: 0.20-0.25%). The highest risk of 4.35% (95% CI: 3.84-4.61%) was estimated for individuals aged 80 years or older, whereas this value was approximately 0 in the youngest age groups (e.g., 3-17 years and 18-39 years). For individuals aged 80 years or above, the adjusted ISR in the vaccinated group (1.25%, 95% CI: 1.11-1.33%) was significantly lower than that in the unvaccinated group (4.77%, 95% CI: 4.21-5.05%) **(Figure 3B)**. The adjusted sCFR and sCSR were 0.96% (95% CI: 0.84-1.01%) and 3.06% (95% CI: 2.70-3.24%), respectively.

**Figure 3.**
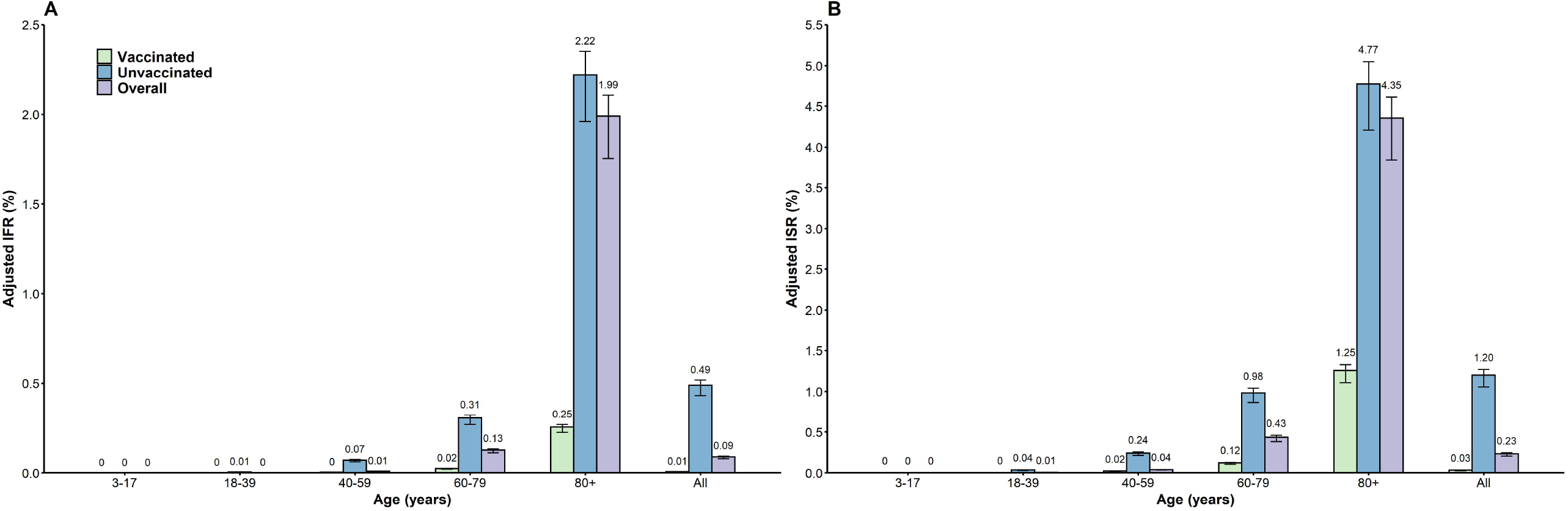
Adjusted IFR and ISR of COVID-19 caused by SARS-CoV-2 Omicron in Shanghai. **A)** IFR; **B)** ISR. The number represents the median estimates, and the error bar represents the 95% confidence interval.

## Discussion

We estimated the disease burden and clinical severity of the SARS-CoV-2 Omicron BA.2 outbreak in Shanghai that occurred between February 26 and June 30, 2022. We estimated that the infection rate, severe/critical infection rate, and mortality rate were 2.74 (95% CI: 2.73-2.74) per 100 individuals, 6.34 (95% CI: 6.02-6.66) per 100,000 individuals and 2.42 (95% CI: 2.23-2.62) per 100,000 individuals, respectively. The severe/critical infection rate and mortality rate were both significantly higher in the elderly population compared with the remainder of the population, and these values were approximately 0 among young individuals (18 years or below). The overall IFR and ISR were 0.09% (95% CI: 0.08-0.10%) and 0.23% (95% CI: 0.20-0.25%), respectively. Having received at least one vaccine dose led to a 10-fold reduction in the risk of death for infected individuals aged 80 years or above.

The Omicron BA.2 outbreak in Shanghai had an intensive community-level transmission, requiring the implementation of strict NPIs. The outbreak was brought under control through a city-level lockdown, traffic shutdown, school closure, repeated and extensive nucleic screenings and medical assistance from other provinces.^9^ This policy minimized the disease burden and number of fatalities. In fact, compared to the disease burden of COVID-19 caused by the Omicron variant in other countries, the overall infection rate and mortality rate in Shanghai were much lower. For example, the infection rate in Shanghai was 2.74 per 100 individuals compared to 34.37 per 100 individuals in South Korea, 13.67 per 100 individuals in Spain, and 24.13 per 100 individuals in Germany. However, in these countries, the number of infected individuals was largely underestimated because population mass screenings were not adopted.^10^ A study in Gauteng, South Africa estimated that the reported deaths and excess deaths associated with COVID-19 were 7 and 12 per 100,000 individuals, respectively, during the Omicron-circulating wave, which are remarkably increased compared with the estimates of 2.42 per 100,000 individuals in Shanghai despite the younger South African population.^11^ Since January 2022, Hong Kong has also experienced a large-scale epidemic caused by the Omicron BA.2 variant. As of June 30, 2022, greater than 1,233,166 infections and 9,188 deaths have been reported by the Hong Kong Health Agency, corresponding to infection and mortality rates of 1,665.0 per 10,000 individuals and 124.1 per 100,000 individuals, respectively.^3,12,13^ The same Omicron variant caused both the Shanghai and Hong Kong outbreaks, and the remarkable difference in mortality could likely be ascribable to the difference in the adopted control measures. In fact, in contrast with Shanghai, the interventions adopted in Hong Kong were less strict and did not aim at the containment of the outbreak. Our previous study showed that if an Omicron epidemic is left untreated (and no spontaneous behaviour change was adopted by the population in response to the epidemic), the number of estimated deaths in mainland China could reach approximately 1.5 million deaths (i.e., approximately 100 per 100,000 individuals - in line with what was observed in the epidemic in Hong Kong).^14^ This estimation further supports the key role that NPIs have played in reducing COVID-19 burden and mortality.

We found a high proportion of asymptomatic infections in Shanghai (90.7%, 95% CI: 90. 7-90.8%), which was greater than that estimated for the ancestral lineage (∼69%)^15^ but consistent with other estimates for other SARS-CoV-2 variants in the presence of vaccination (∼85%).^16^ Lower proportions of asymptomatic infections were reported in previous studies,^17^ but those estimates were obtained in the absence of repeated screenings of the population. Moreover, it is important to note that the criteria adopted for the definition of asymptomatic infection may vary across locations and study periods. We estimated the overall IFR of the Omicron BA.2 outbreak in Shanghai to be 0.09% (95% CI: 0.08-0.10%), which is less than that of Hong Kong (a modelling study estimated an IFR of 0.19%).^18^ This difference may be explained by the shortage of hospital beds experienced by Hong Kong during the Omicron wave^18^ and differences in vaccination coverage. Vaccination coverage for the primary and booster doses is higher in Shanghai than in Hong Kong, especially among people aged 65 years or older (coverage of booster: 38% in Shanghai vs. 18% in Hong Kong before the Omicron outbreak).^12^ Among deceased cases reported in Hong Kong, greater than 70% of cases were never vaccinated.^3^ Although the vaccines used in these two cities are not the same (inactivated vaccine in Shanghai, both inactivated and mRNA vaccines in Hong Kong), several studies have shown that the effectiveness against severe/critical COVID-19 disease or death is not significantly different between these two different platforms.^19^ More broadly, it is important to stress that increasing vaccine coverage could effectively reduce the clinical severity of COVID-19. We comprehensively summarized estimates of IFR as well as local vaccine-coverage data for several different settings, and we found that the IFR was lower in the setting with higher coverage of primary or booster vaccination, regardless of the type of circulating SARS-CoV-2 **(Appendix p2, p8-10)**. However, this analysis did not control for the different levels of natural infection and surveillance capacity in the study locations; this would require the collection of additional data, which are not readily available to us. Increasing vaccination coverage in Shanghai, especially for older people with a history of underlying chronic diseases, and overcoming vaccine hesitancy are critically important in preparation for possible future waves of COVID-19.

Our study has several limitations. First, due to the lack of reliable vaccination data, we could not estimate COVID-19 burden by vaccination status. Second, some asymptomatic infections who developed very mild symptom were not sent to the designated isolation hospital and were not counted as symptomatic cases, resulting in possible misclassification bias. Third, two types of RT□PCR kits were used for case detection and massive nucleic acid screening during the outbreak (BioGerm and Bioperfectus); the actual usage and distribution of these two kits is unknown. Thus, we used RT□PCR sensitivity from a single manufacturer (BioGerm) in the main analysis. However, we conducted a sensitivity analysis using the sensitivity estimated for the other kit (Bioperfectus) and found no significant difference compared to the main results.

In conclusion, we estimated the disease burden and clinical severity of the Omicron BA.2 outbreak in Shanghai in February-June 2022. We found a lower burden and mortality of the outbreak compared to other settings and countries, showing the impact of successful outbreak containment in Shanghai. The estimated low clinical severity of this Omicron BA.2 epidemic in Shanghai highlighted the key contribution of vaccination and availability of hospital beds to reduce the risk of death.

## Data Availability

The data and code that support the findings of this study will be made available in GitHub upon manuscript acceptance.

## Contributors

H.Y. conceived, designed, and supervised the study. X.C., X.Y., N.Z., R.S., J.Z., W.L. and T.Z. collected and checked the data. X.C., X.Y., J.Z. and N.Z. analysed the data. X.C. wrote the first draft of the manuscript. X.C., K.S., J.C., J.Z., X.D., M.A. and H.Y. interpreted the results and revised the content critically. All authors approved the final version for submission and agreed to be accountable for all aspects of the work.

## Acknowledgements

The findings and conclusions in this report are those of the authors and do not necessarily represent the official position of the NIH. This study was supported by grants from the Key Program of the National Natural Science Foundation of China (grant 82130093 to H.Y.). The funders had no role in the study design, data collection, data analysis, data interpretation or writing of the report.

