## Supplements for "Estimation of disease burden and clinical severity of COVID-19 caused by Omicron BA.2 in Shanghai, February-June 2022"

### Methods and Results

**Correlation analysis between clinical severity and vaccine coverage**

Using predefined search terms (**Table S2**), we conducted a rapid literature review and comprehensively collected studies reporting IFR and/or sCFR in other settings. The summarized results are shown in Table S3. We also extracted vaccine coverage from multiple data sources for the included studies. We explored the impact of vaccine coverage of primary and booster vaccinations associated with IFR and sCFR using a linear regression model. The Pearson correlation coefficient was calculated, and the p value was reported for each correlation analysis. Furthermore, we conducted a correlation analysis by age group.

We found that IFR was apparently lower in the setting with higher coverage of primary or booster vaccination regardless of the type of circulating SARS-CoV-2 **(Figure S6)**. The submicron-specific IFR was also well correlated with the COVID-19 vaccine coverage of the primary series (Pearson correlation coefficient: -0.99; p=0.015) **(Figure S7)**. We further stratified such estimates by age groups by correlating age-specific coverage across different locations, revealing a similar pattern of lower IFR as the coverage rate increased **(Figure S8)**.

### Figure S1 Timeline of nucleic acid screening during the outbreak in Shanghai, Feb-May 2022

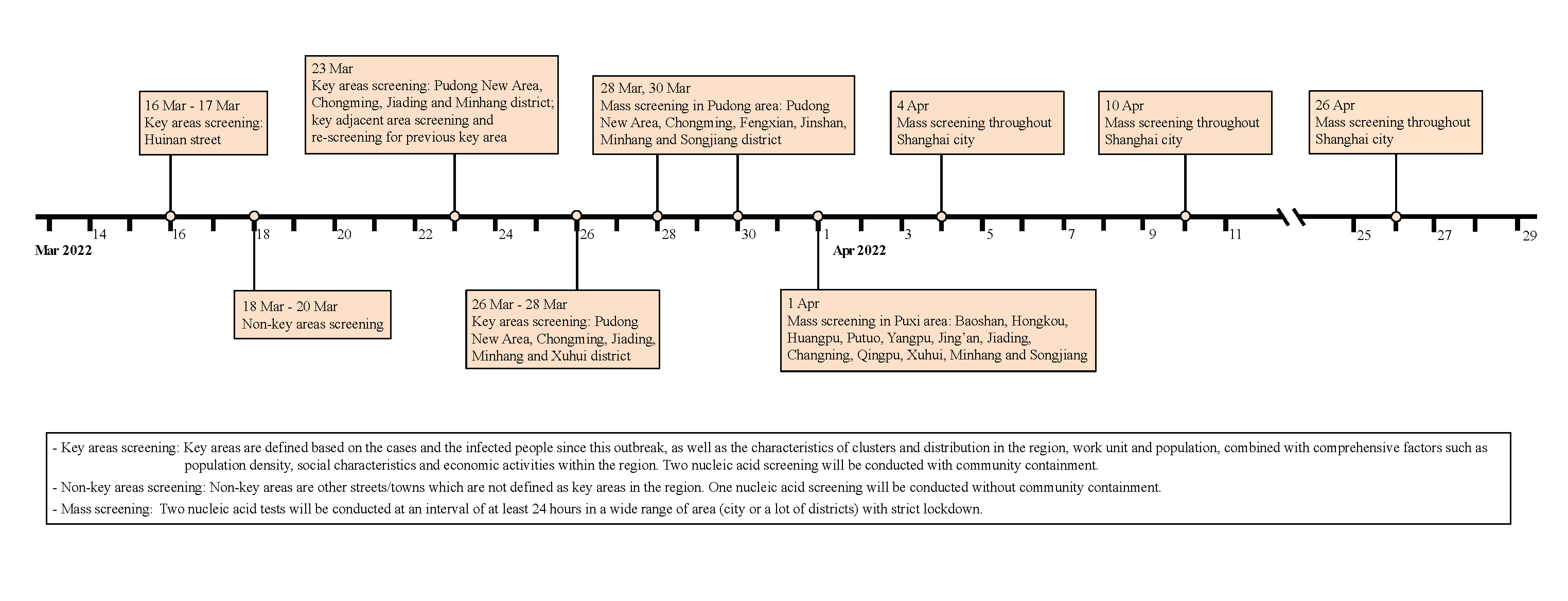

### Figure S2 Asymptomatic proportion of COVID-19 caused by SARS-CoV-2 Omicron BA.2 infection during the Shanghai outbreak

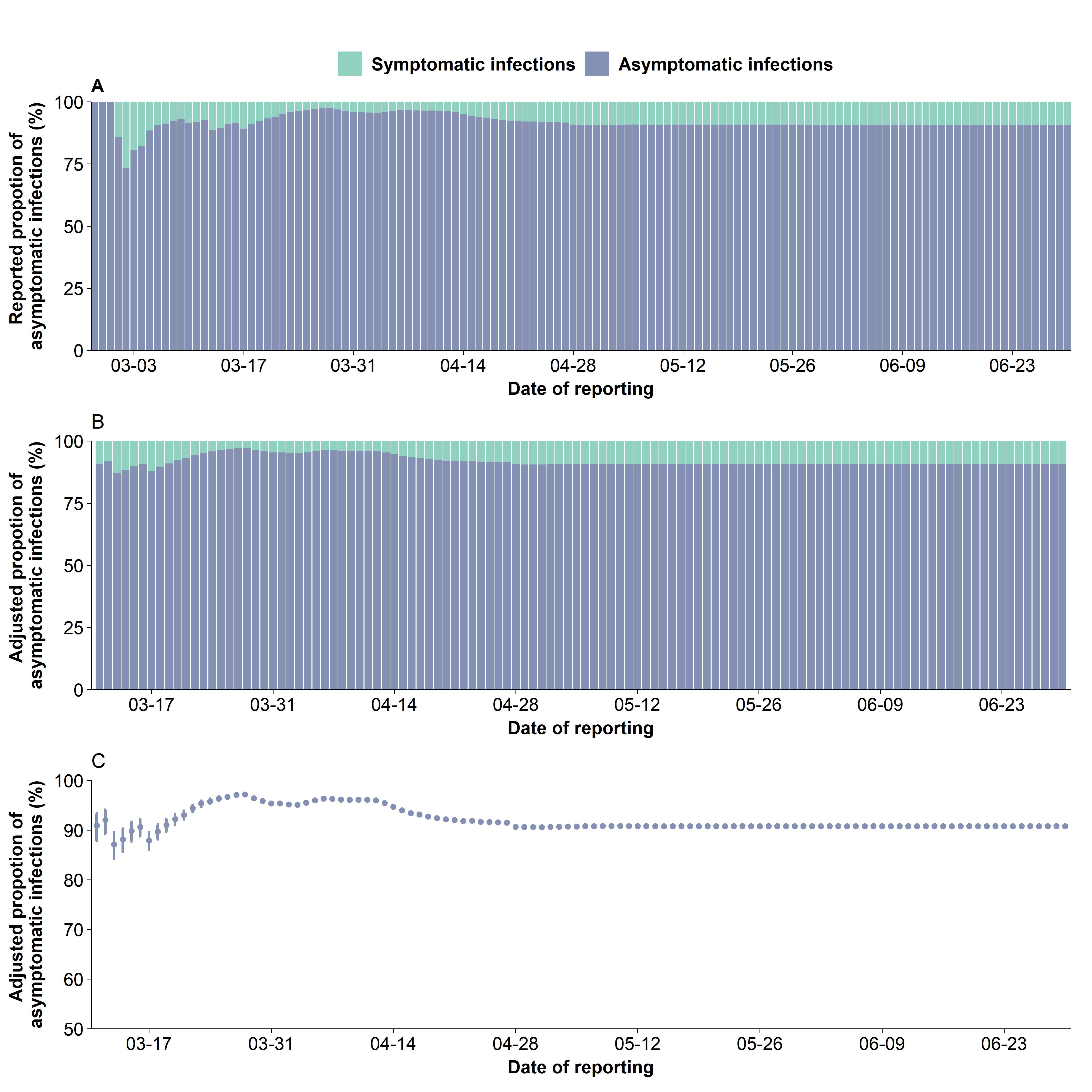

### Figure S3 Time-varying infection case fatality risk of COVID-19 caused by SARS-CoV-2 Omicron BA.2 infection during the Shanghai outbreak

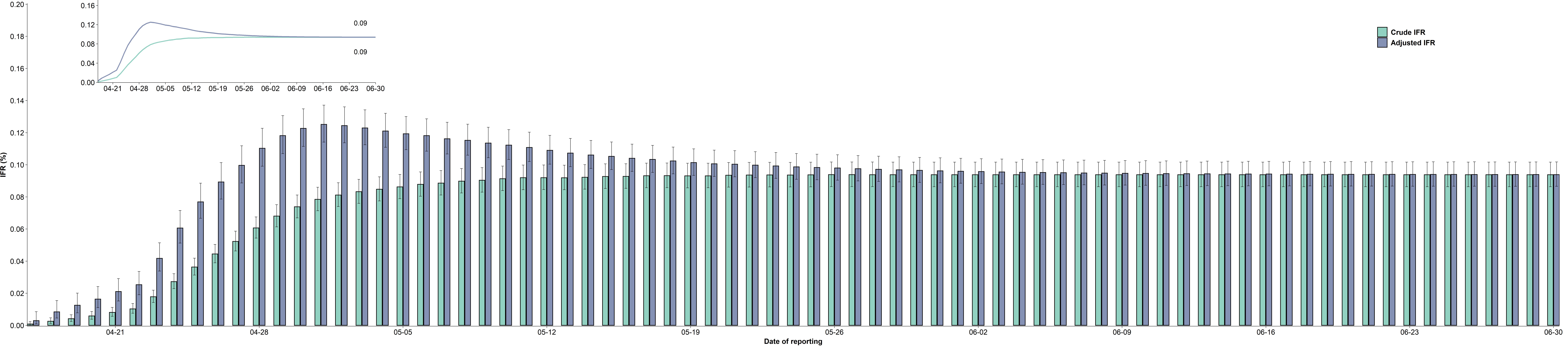

### Figure S4 Sensitivity analysis of the infection rate by changing RT-PCR sensitivity

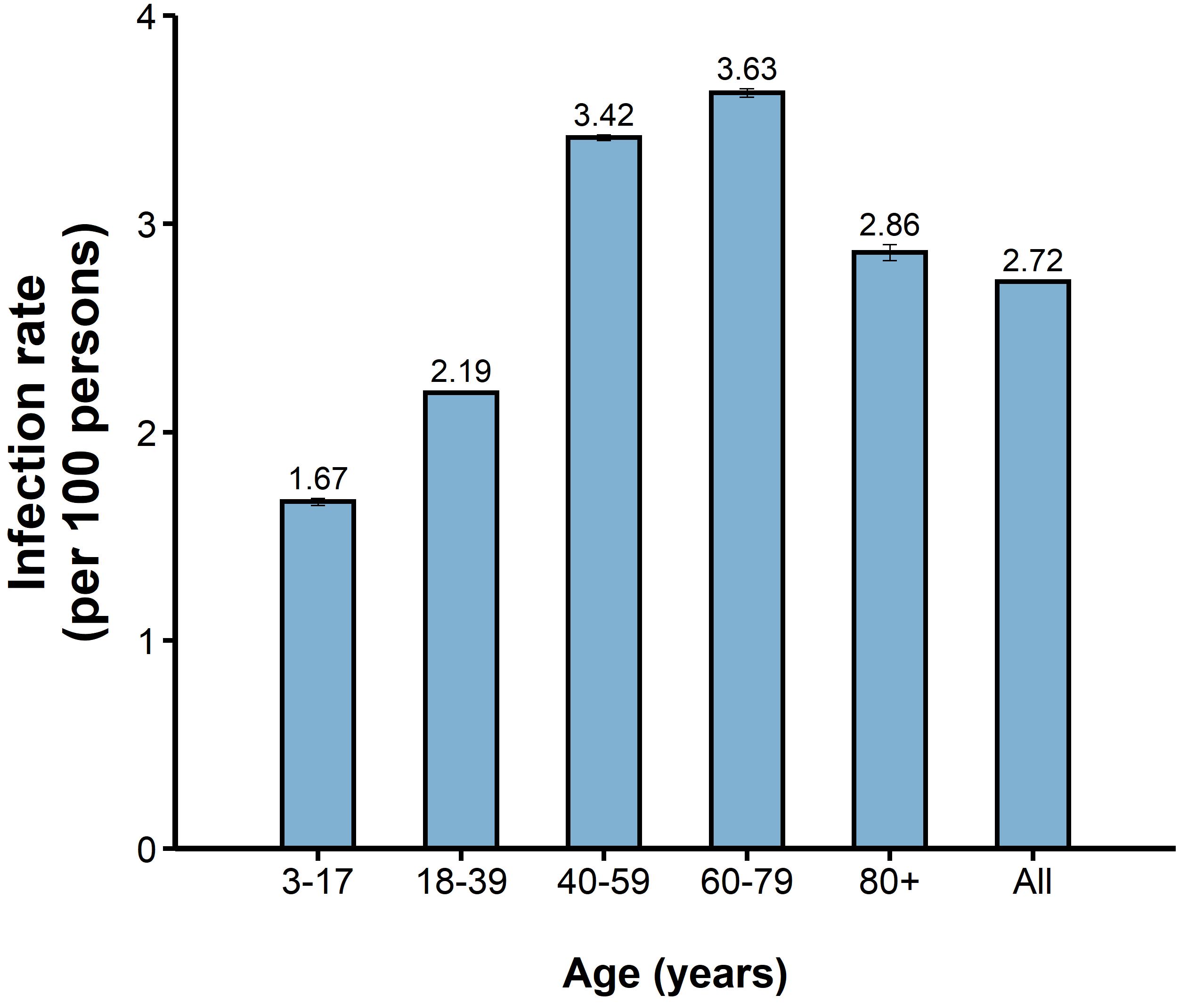

### Figure S5 Sensitivity analysis of IFR and ISR by changing RT-PCR sensitivity

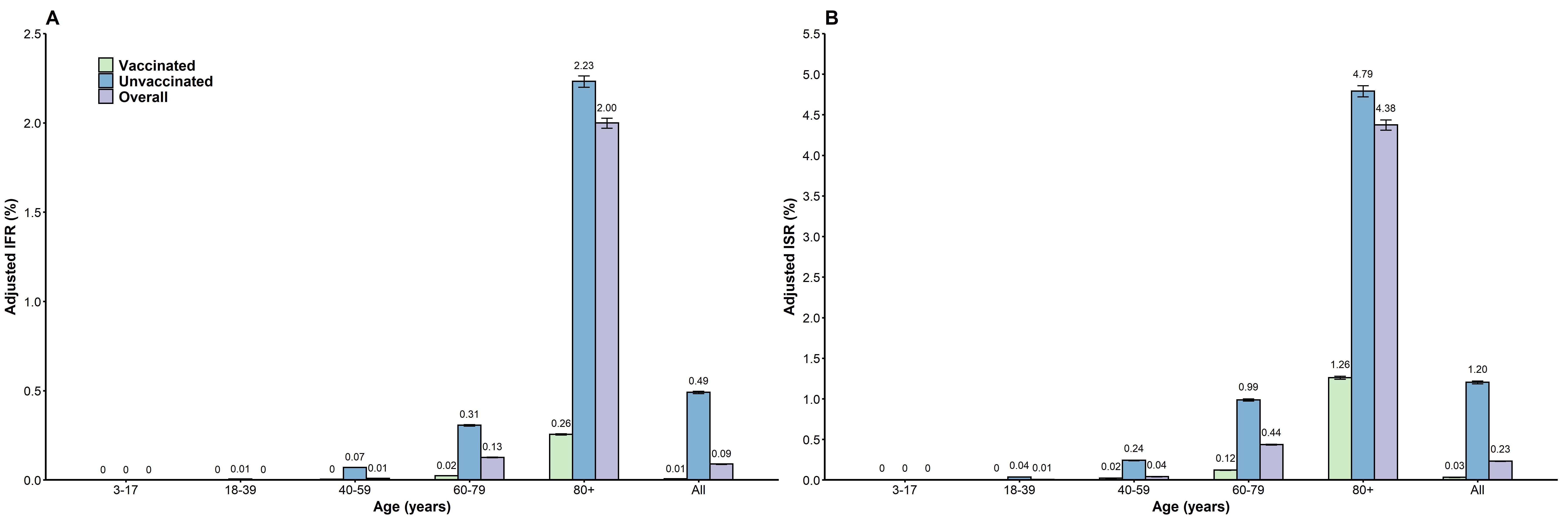

### Figure S6 Correlation analysis between IFR and vaccine coverage

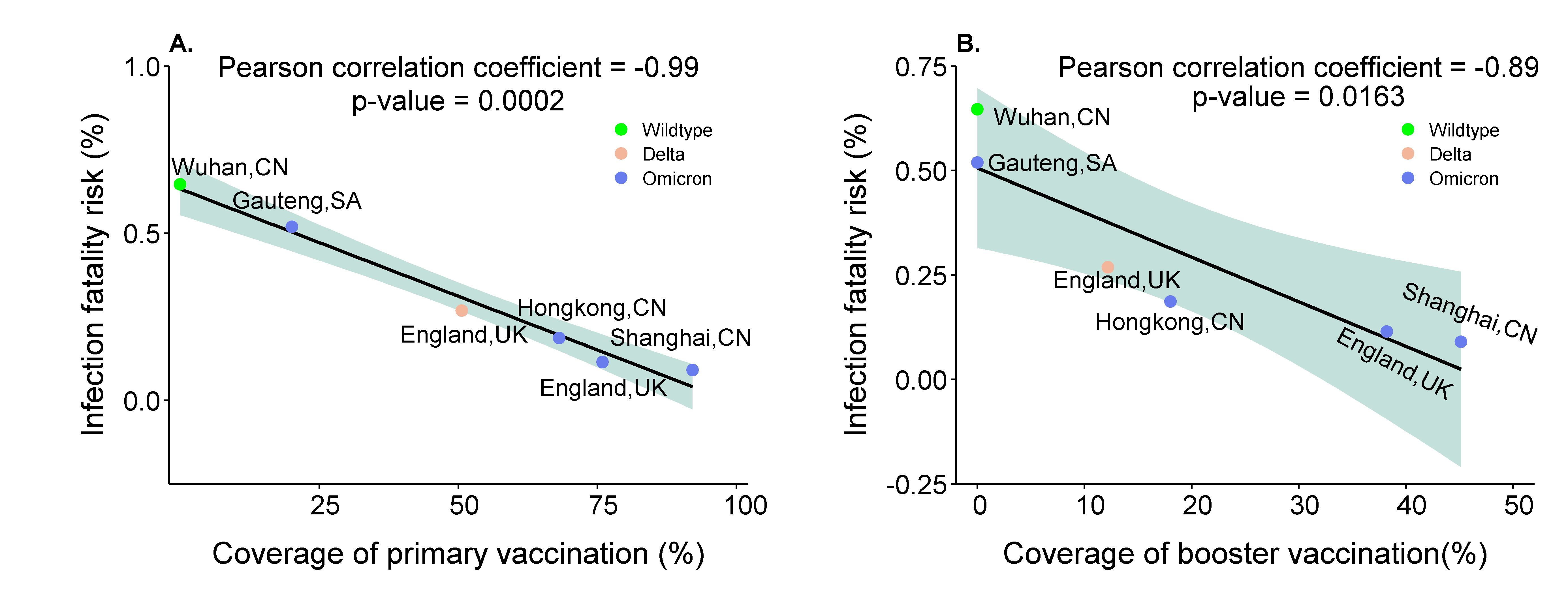

### Figure S7 Correlation analysis between Omicron-specific IFR and vaccine coverage

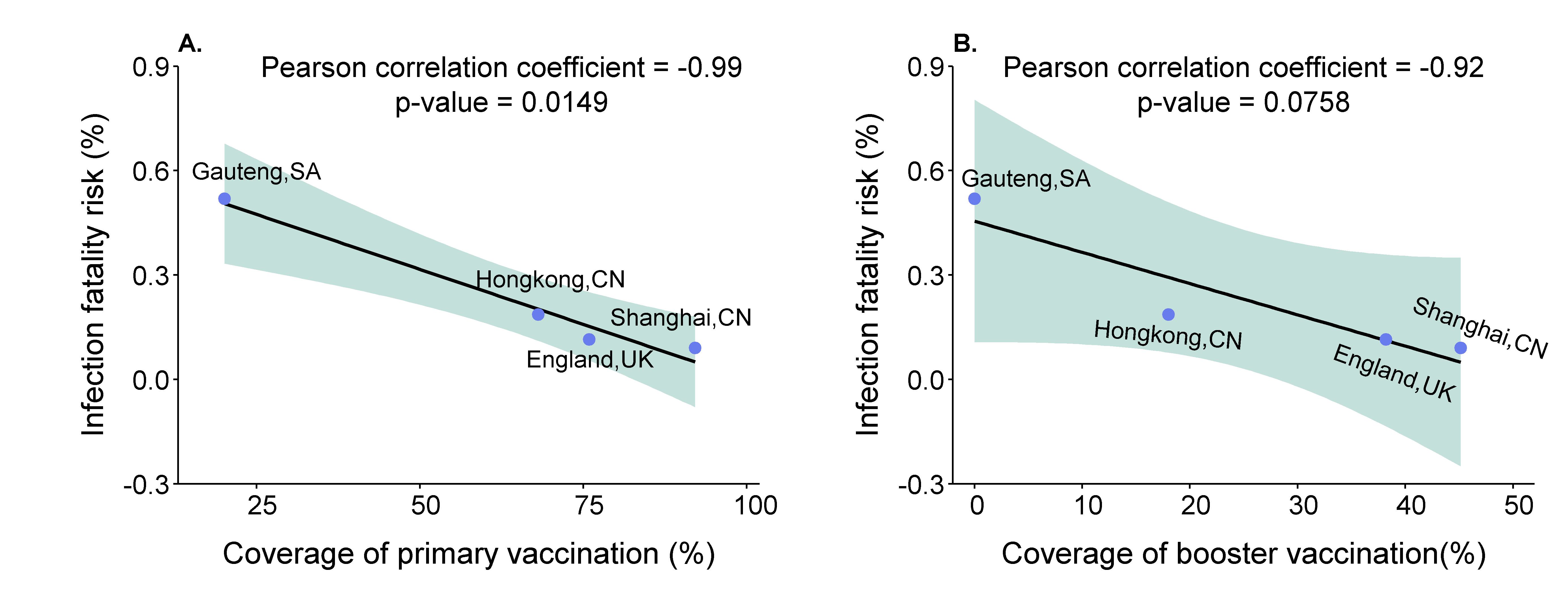

### Figure S8 Correlation analysis between Omicron-specific IFR and vaccine coverage by age group

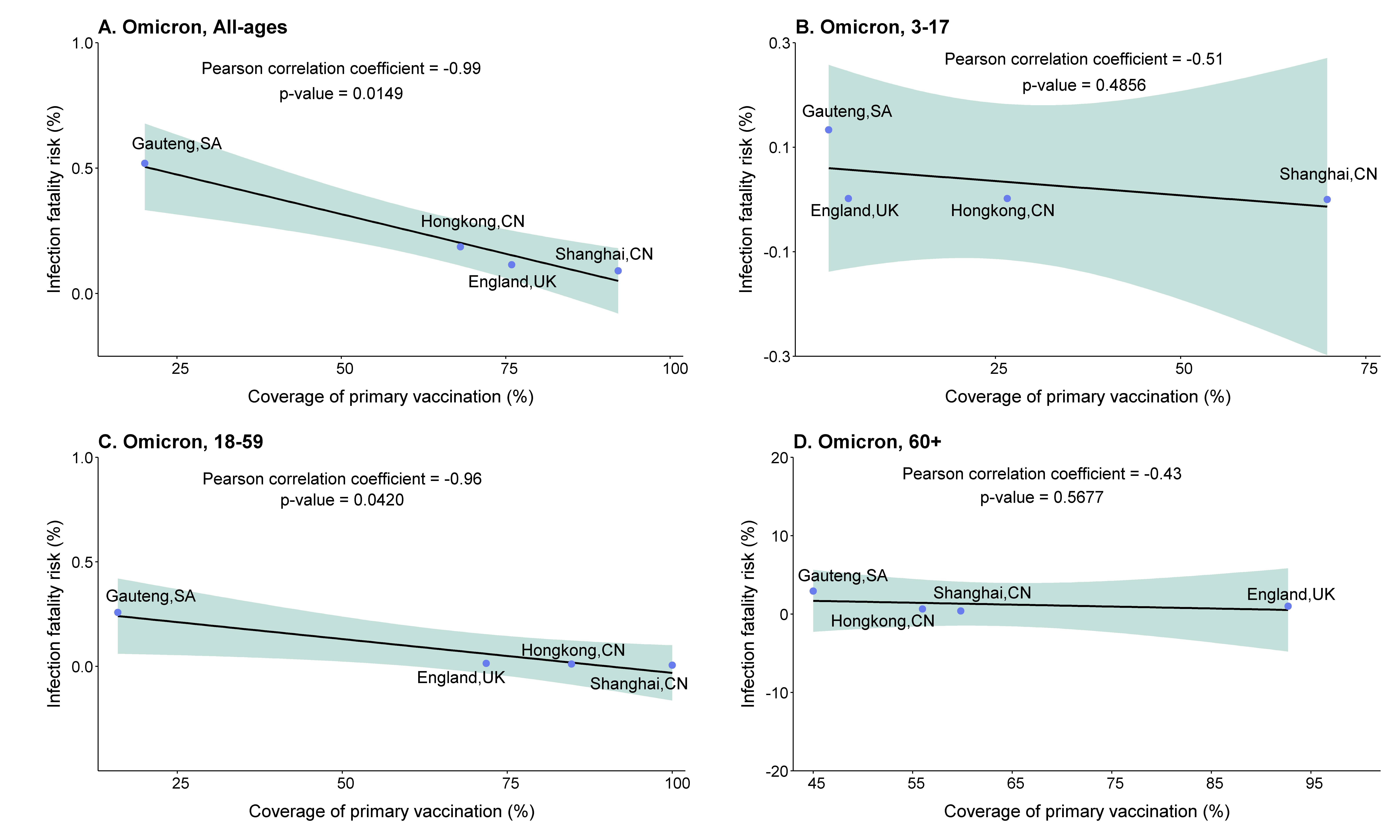

### Table S1 Data sources and parameters used in the study

| **Variable** | **Definition** | **Sources** |
| --- | --- | --- |
| **Data** |  |  |
| Individual-data line-list | Individual data includes age, address, whether symptomatic, and whether changing status from asymptomatic to symptomatic infections | The official websites of Shanghai municipal Health Commissions  The official websites of Shanghai municipal Health Commissions^1^ |
| Number of reported confirmed cases | Cumulative number of PCR-confirmed infections, including both symptomatic and asymptomatic case, by report date since Feb 26, 2022. |  |
| Number of reported symptomatic cases | Cumulative number of PCR-confirmed symptomatic infections by report date since Feb 26, 2022. |  |
| Number of reported deaths | Cumulative number of deceased cases by report date since Feb 26, 2022. |  |
| Population size | Shanghai resident population at the end of 2020 | Shanghai Statistical Yearbook 2021^2^ |
| **Parameters** |  |  |
| Age distribution of deaths | Before May 15, 2022, the age distribution of 568 deceased cases were extracted from the literature. The age of daily new deceased cases was reported by Shanghai Municipal Health Commissions since then. | The official websites of Shanghai municipal Health Commissions^1^;  Wang et al., personal communication |
| Age distribution of infections | Age distribution of 612,597 infections from Dec 2, 2021 and May 13, 2022 was extracted from the literature. | Wang et al., personal communication |
| Age distribution of severe/critical patients | Age distribution of 1485 severe/critical patients from Dec 2, 2021 and May 13, 2022 was extracted from the literature. | Wang et al., personal communication |
| Vaccination status of deaths | Vaccination status of 612,597 infections cases was extracted from the literature. | Wang et al., personal communication |
| Vaccination status of infections | Vaccination status of 1485 severe/critical patients was extracted from the literature. | Wang et al., personal communication |
| Vaccination status of severe/critical patients | Vaccination status of 568 deceased cases was extracted from the literature. | Wang et al., personal communication |
| Sensitivity of the RT‒PCR | BioGerm: 0.944 (95%CI: 0.706-0.997); | Lu et al., 2020^3^ |
| Interval between infection and symptom onset | 1.91 days (95% CI: 0.25-14.28) with lognormal distribution | Yu et al., unpublished data |
| Interval between symptom onset and admission | 0.92 days (95% CI: 0.13-6.61) with lognormal distribution | Yu et al., unpublished data |
| Interval between admission and death | 6.92 days (95% CI: 0.53-29.52) with gamma distribution | Yu et al., unpublished data |
| Interval between infection and severe/critical illness | 5.7 days (95% CI: 4.1-7.8) with lognormal distribution | Yu et al., unpublished data |

### Table S2 Search strategy and terms

| **Database** | **Step** | **Search strategy** |
| --- | --- | --- |
| PubMed | #1 | Title/Abstract: 2019-nCoV OR “coronavirus disease 2019” OR COVID-19 OR “severe acute respiratory syndrome coronavirus 2” OR SARS-CoV-2 |
|  | #2 | MeSH Terms: SARS-CoV-2 OR COVID-19 |
|  | #3 | #1 OR #2 |
|  | #4 | Title/Abstract: VOC OR “Variant of Concern” OR “Variants of Concern” OR variant* OR mutant* OR mutation* OR “Omicron” OR “B.1.1.529” OR “GR/484A” OR “21K” OR “21L” OR “21M” OR “BA.1” OR “BA.2” OR “BA” OR “BA.2.12.1” |
|  | #5 | Title/Abstract: IFR* OR serology confirmed infection fatality risk OR infection fatality risk OR mortality OR fatality OR death OR case fatality risk OR CFR OR symptomatic case fatality risk OR sCFR |
|  | #6 | Language: English |
|  | #7 | Nov 1, 2021 to June 30, 2022 |
|  | #8 | #3 AND #4 AND #5 AND #6 AND #7 |
| Web of Science | #1 | Title/Abstract: 2019-nCoV OR “coronavirus disease 2019” OR COVID-19 OR “severe acute respiratory syndrome coronavirus 2” OR SARS-CoV-2 |
|  | #2 | Title/Abstract: VOC OR “Variant of Concern” OR “Variants of Concern” OR variant* OR mutant* OR mutation* OR “Omicron” OR “B.1.1.529” OR “GR/484A” OR “21K” OR “21L” OR “21M” OR “BA.1” OR “BA.2” OR “BA” OR “BA.2.12.1” |
|  | #3 | Title/Abstract: IFR* OR serology confirmed infection fatality risk OR infection fatality risk OR mortality OR fatality OR death OR case fatality risk OR CFR OR symptomatic case fatality risk OR sCFR |
|  | #4 | Language: English |
|  | #5 | 2021/11/01 to 2022/04/25 |
|  | #6 | #1 AND #2 AND #3 AND #4 AND #5 |
| Embase | #1 | Title/Abstract: 2019-nCoV OR “coronavirus disease 2019” OR COVID-19 OR “severe acute respiratory syndrome coronavirus 2” OR SARS-CoV-2 |
|  | #2 | Title/Abstract: VOC OR “Variant of Concern” OR “Variants of Concern” OR variant* OR mutant* OR mutation* OR “Omicron” OR “B.1.1.529” OR “GR/484A” OR “21K” OR “21 L” OR “21 M” OR “BA.1” OR “BA.2” OR “BA” OR “BA.2.12.1” |
|  | #3 | Title/Abstract: IFR* OR serology confirmed infection fatality risk OR infection fatality risk OR mortality OR fatality OR death OR case fatality risk OR CFR OR symptomatic case fatality risk OR sCFR |
|  | #4 | Language: English |
|  | #5 | 2021/11/01 to 2022/04/25 |
|  | #6 | #1 AND #2 AND #3 AND #4 AND #5 |
| EuropePMC | #1 | Title/Abstract: 2019-nCoV OR “coronavirus disease 2019” OR COVID-19 OR “severe acute respiratory syndrome coronavirus 2” OR SARS-CoV-2 |
|  | #2 | Title/Abstract: VOC OR “Variant of Concern” OR “Variants of Concern” OR variant* OR mutant* OR mutation* OR “Omicron” OR “B.1.1.529” OR “GR/484A” OR “21K” OR “21 L” OR “21 M” OR “BA.1” OR “BA.2” OR “BA” OR “BA.2.12.1” |
|  | #3 | Title/Abstract: IFR* OR serology confirmed infection fatality risk OR infection fatality risk OR mortality OR fatality OR death OR case fatality risk OR CFR OR symptomatic case fatality risk OR sCFR |
|  | #4 | Language: English |
|  | #5 | 2021/11/01 to 2022/04/25 |
|  | #6 | #1 AND #2 AND #3 AND #4 AND #5 |

### Table S3 Summary of infection case fatality risk of COVID-19 caused by Omicron infection

| **Authors (year)** | **Location** | **Time** | **Variant** | **Methods** | **Case definition** | **Data sources/Case identification** | **Study participants** | **Results** |
| --- | --- | --- | --- | --- | --- | --- | --- | --- |
| Infection-fatality risk |  |  |  |  |  |  |  |  |
| - | Wuhan, China | Dec 2019-Mar 2020 | Prototype | Using age-specific serology data in Wuhan combined with data from the total Wuhan general population, the number of serology-confirmed infections was calculated. We further estimated crude infection fatality risk by dividing by the number of deceased cases across the corresponding age groups. | - | **Seroprevalence by age group in Wuhan (%)^5^**  0-5 years: 14/303 (4.6);  6-11 years: 23/682 (3.4)  12-17 years: 16/485 (3.3)  18-44 years: 214/3905 (5.5)  45-65 years: 202/3340 (6.0)  ≥66 years: 63/827 (7.6)  **Combined with total population in Wuhan**  Overall: 10696970  0-19 years: 2002195  20-39 years: 3885005  40-59 years: 3363579  ≥60 years: 1446191  **Cumulative deceased cases by age group in Wuhan (%)^6^**  Overall: 3869  0-19 years: 4  20-39 years: 95  40-59 years: 635  ≥60 years: 3135 | - | **Estimated infections by age group**  Overall: 1507661  0-17 years: 385673  18-65 years: 307104  ≥66 years: 814884  **Estimated IFR by age group (%)**  Overall: 3869/1507661 (0.26)  0-19 years: 4/385673 (0.001)  20-59 years: 730/307104 (0.24)  ≥60 years: 3135/814886 (0.38) |
| Nyberg et al., Lancet (2022)^7^ | England, UK | Nov 2021-Jan 2022 | Omicron and Delta | The study adopted individual-level data based on laboratory-confirmed cases linked to routine datasets on vaccination status, hospital attendance and admission, and mortality. The relative risk of hospital attendance or admission within 14 days or death within 28 days after confirmed infection was estimated using proportional hazards regression. | Individual resident in England with laboratory-confirmed (whole-genome sequencing confirmed/genotyping-confirmed/S gene positivity) SARS-CoV-2 infection  The mortality outcome was defined as death occurring 0–28 days after the first positive specimen date of the most recent infection episode, again matching the definition used in routine UK government reporting. | All positive cases in England are reported to the UK Health Security Agency (UKHSA), which maintains a definitive **line** **list of all individuals who have had confirmed SARS-CoV-2 infection**. The surveillance activities within which this study was conducted are part of UKHSA’s responsibility to monitor COVID-19 during the current pandemic.  The sequencing/genotyping strategy includes geographically weighted, population-level sampling of community cases, supplemented by targeted sampling of recent international travellers, hospitalized cases, and hospital staff. | A total of 1,516,702 cases were included, including 1,067,859 Omicron-infected cases and 448,843 Delta-infected cases. Approximately 75% cases have been vaccinated with at least one dose of AstraZeneca/Pfizer/Moderna COVID-19 vaccine, 68.5% cases have been vaccinated with at least two doses of AstraZeneca/Pfizer/Moderna COVID-19 vaccine, and 30% cases have been vaccinated with 3 doses of AstraZeneca/Pfizer/Moderna COVID-19 vaccine. | **Omicron (%)**  Overall: 1225/1067859 (0.11)  <10 years: 2/43147 (0.005)  10-19 years: 1/119261 (0.001)  20-29 years: 5/265199 (0.002)  30-39 years: 11/228915 (0.005)  40-49 years: 25/167045 (0.01)  50-59 years: 71/134186 (0.05)  60-69 years: 128/64875 (0.20)  70-79 years: 257/31066 (0.83)  ≥80 years: 725/14165 (5.12)  **Delta (%)**  Overall: 1205/448843 (0.27)  <10 years: 1/74484 (0.001)  10-19 years: 1/88284 (0.001)  20-29 years: 2/48178 (0.004)  30-39 years: 28/79401 (0.04)  40-49 years: 43/81760 (0.05)  50-59 years: 128/49298 (0.26)  60-69 years: 206/18330 (1.12)  70-79 years: 294/5943 (4.95)  ≥80 years: 502/3165 (15.9) |
| Madhi et al., NEJM (2022)^8^ | Gauteng, South Africa | Mar 2021- | Delta, Omicron | The study evaluated Covid-19 epidemiologic trends in Gauteng, including cases, hospitalizations,  recorded deaths, and excess deaths, from the start of the pandemic through January 12, 2022. | Cases included asymptomatic and symptomatic infections with SARS-CoV-2 confirmed by either a nucleic acid amplification assay or a rapid antigen test. | Data regarding daily cases, hospitalizations, and  recorded deaths were sourced from the South  African National Institute for Communicable  Diseases daily databases, including the DATCOV  database, through January 12, 2022. | - | **Omicron (%)**  0-4 years: 19/4724 (0.40)  5-19 years: 22/26067 (0.08)  20-49 years: 299/147270 (0.20)  50-59 years:146/25408 (0.57)  >60 years: 692/23462 (2.95)  **Delta (%)**  0-4 years: 49/6709 (0.73)  5-19 years: 51/53636 (0.10)  20-49 years: 2566/300032 (0.86)  50-59 years: 3144/81120 (3.88)  >60 years: 8873/70142 (12.65) |
| HKU WHO Collaborating Centre on Infectious Disease Epidemiology and Modelling (2022)^9^ | Hongkong, China | Jan-Mar, 2022 | Omicron | Using modelling analysis, the epidemic size of the fifth wave by April 21 was predicted to be approximately 4.5 million (CrI: 4.4-4.6million) infections and 8,383 (CrI: 7,588–9,241) deaths if antiviral supply is sufficient and liberally deployed. Combined with age distribution of reported cases and death from Centre for Health Protection of the Department of Health and the Hospital Authority, age-specific infection fatality risk could be estimated.^10^ | - | - | - | **Crude infection fatality risk (%)**  Overall: 8383/4500000 (0.18)  0-17 years: 8/463500 (0.002)  18-59 years: 319/2790000 (0.01)  ≥60 years: 8056/1246500 (0.65) |
| Case fatality risk |  |  |  |  |  |  |  |  |
| Yang et al., Nature Communication (2020)^6^ | Wuhan, China | Dec 2019-Mar 2020 | Prototype | Using multiple data sources, the number of symptomatic cases during the first wave of Wuhan outbreaks were estimated by adjusting both PCR sensitivity and health care-seeking behaviours. | Cases include RT‒PCR confirmed cases and clinically diagnosed cases, which were comply to the diagnosis and treatment scheme of novel coronavirus diseases 2019 initiated by National Health Commission of China. | Reported number of cases were extracted from Wuhan Municipal Health Commission; other parameters were retrieved from published paper. | - | Overall: 4.54 (3.70-5.14)  0-17 years: 0.36 (0.33-0.40)  18-59 years: 1.28 (1.18-1.37)  ≥60 years: 9.09 (6.80-10.59) |
| Christensen et al., medRxiv  (2022)^11^ | Houston, USA | Nov 2021-Dec 2021 | Omicron, Delta | For analyses comparing features of patients infected with the Omicron VOC and Alpha and Delta VOCs, all patients documented to be infected with these variants in the Houston Methodist system were studied. | Cases were confirmed by molecular diagnostic testing, genome sequencing and S-gene target-failure (SGTF), and sequencing. The great majority of individuals had signs or symptoms consistent with COVID-19 disease. | Data and specimens were obtained from patients registered at Houston Methodist facilities (e.g., hospitals and urgent care centres) and institutions in the Houston metropolitan region. | **-** | **Omicron:** 38/4468 =0.9%  **Delta:** 839/15728 = 5.3% |
| Davies et al., medRxiv  (2022)^12^ | South Africa | Nov 2021-Dec 2021 | Omicron, Delta | This cohort study compared the risk between waves of the outcomes, including death and severe hospitalization, using Cox regression of data from the Western Cape Provincial Health Data Centre (WCPHDC). | Public sector **patients** aged ≥20 years with a laboratory confirmed COVID-19 diagnosis (positive SARS-CoV-2 PCR or antigen test) during the 4^th^ omicron wave.  Deaths were defined as deceased cases within 14 days of COVID-19 diagnosis. | Deidentified data were collected from WCPHDC, which consolidates administrative, laboratory, and pharmacy data from routine electronic clinical information systems used in all public sector health facilities with linkage through a unique identifier. | A total of 5,104 Omicron-infected cases in the fourth wave and 4355 Delta-infected cases in the third wave were included. In total, 38% cases were fully vaccinated during the Omicron wave, and 2.8% of cases were fully vaccinated during the Delta wave. | Omicron: 20/5104=0.4%  Delta: 250/4355=5.7% |
